# Evaluation of applicability of the online version of HADS-D for depression phenotype screening in the general population

**DOI:** 10.1101/2020.10.16.20213843

**Authors:** AA Kibitov, AS Rakitko, ED Kasyanov, GV Rukavishnikov, KA Kozlova, VV Ilinsky, NG Neznanov, GE Mazo, AO Kibitov

## Abstract

One of the most promising areas of research into the biological underpinnings of depression is genetic studies. However, the absence of generally accepted phenotyping methods leads to the difficulties in generalizing their results due to the heterogeneity of the samples. Thus, the development of a reliable and convenient phenotyping method that allows large sample sizes to be included in studies remains a top priority for the further development of genetic studies of depression.

The aim of this study was to evaluate the applicability of the online version of the depression subscale of Hospital Anxiety and Depression Scale (HADS-D) for depression phenotype screening in the general population. Using online HADS-D we performed screening of depressive symptoms and compared results with known population patterns of depression.

We conducted an online survey of 2610 Russian-speaking respondents over the age of 18. The overall HADS-D score was higher in women (p=0.003), in individuals under 30 y.o compared to participants over 42 y.o. (p=0.004) and in individuals reporting cardiovascular diseases (CVD) symptoms (p<0.0001).

Linear regression showed that the presence of CVD leads to higher HADS-D scores (p<0.001), male gender (p=0.002) and older age (p<0.001) led to lower scores. Logistic regression showed that CVD increases the risk of having depression symptoms by HADS-D (p=0.033, OR=1.29), older age (p=0.015, OR=0.87) and male sex (as a trend, p=0.052, OR=0.80) decrease this risk.

These results are consistent with the known data on the association of sex, age, and the presence of CVD with the prevalence of depression. The online version of HADS-D, given the ease of its usage, can be regarded as an effective tool for phenotyping depression in the general population.

---

Depression is one of the most common mental disorders. According to the World Health Organization, depression is one of the leading causes of disability and contributes significantly to the global burden of disease [1]. Given the extreme social significance of depression, the study of its biological underpinnings is considered to be one of the priority areas in medical research.

To date, a large amount of data has been accumulated on the significant role of genetic factors in depression risk and development. That makes it possible to regard genetic studies as a promising direction for studying its biological mechanisms and identifying biomarkers of the risk [2]. Today, the most informative and promising field of research are genome-wide association studies (GWAS), which require the largest possible sample size and qualitative phenotypic criteria while maintaining a reasonable ratio of research cost and the value of its potential result [3].

A number of GWAS studies on depression have been conducted recently, but generalizing their results is still difficult, mainly due to the extreme heterogeneity of the samples in these studies. This variability is caused both by the clinical heterogeneity of depression and by the use of different approaches to depression phenotyping, which is a consequence of the absence of generally accepted clinical phenotypes of this condition [2].

In some works, depression is determined on the basis of criteria specified in modern diagnostic classifications (ICD (International Classification of Diseases) and DSM (Diagnostic and Statistical Manual of Mental Disorders) of various revisions), which requires a personal contact with a psychiatrist during a clinical interview. This approach is difficult when analyzing large samples due to the high costs and high expending of resources [4]. Other works use a variety of diagnostic scales and questionnaires for self-completion (including online versions), medical records and registers [5]. Finally, in some works “minimal phenotyping” is used, based on self-reports of respondents about the presence or absence of a diagnosis of depression or the fact of taking antidepressants. This approach allows researchers to analyze very large samples (up to hundreds of thousands of participants), but the quality of phenotyping is significantly deteriorating [6].

Thus, the development of reliable and convenient methods for depression phenotyping in the general population remains a top priority for the further development of genetic studies of depression. This will allow unifying the approach and increasing the homogeneity of samples. An important condition for such a method should be the ability to analyze large samples and relative simplicity while maintaining clinical validity.

One of the possible options is using the screening psychometric scales. Their use in research practice has a number of advantages. First, the screening scales are self-questioning, which can simplify data collection and increase sample sizes. Second, the screening scales are based on a continuous quantitative approach, which provides additional opportunities for analyzing not only categorical, but also quantitative elements of the depression phenotype, increasing the quality of phenotyping. Nevertheless, remains the question of their effectiveness and reliability in identifying depression as a clinical phenomenon. This can be assessed by comparing the results of the screening with patterns of depression prevalence in the general population. These include the high prevalence and severity of depression among women [7]; the heterogeneity of the prevalence of depression in different age groups [8]; and pronounced association of depression with cardiovascular diseases (CVD) [9].

One of the tools for depressive symptoms screening in the context of the population patterns mentioned above can be the Hospital Anxiety and Depression Scale (HADS) [10]. The data accumulated over the years of using HADS have shown that this scale is a fairly reliable tool for screening anxiety and depression in general practice patients [11]. HADS consists of two subscales - anxiety and depression. This allows depressive symptoms to be assessed separately using only the corresponding subscale - HADS-D (HADS - Depression). In addition, HADS does not include questions related to autonomic and general somatic symptoms, which can “overlap” with other nosologies and lead to false positive results. In addition, the simple structure of HADS makes it possible to use it in an online questionnaire format, which can simplify the data collection procedure, potentially preserving the accuracy of this tool in its ability to detect depressive disorders in the general population.

There are some publications about the possibilities of using online-HADS-D for assessing depressive symptoms, but only in specific cohorts: in patients with cystic fibrosis [12] and in patients with tinnitus [13]. The potential of the online version of HADS as a screening tool for the detection and phenotyping of depression in the general population has not yet been explored.

Thus, the aim of this study was to investigate the capabilities and applicability of the online version of HADS-D as a potential tool for phenotyping depression in the general population.

## METHODS

The clients of Genotek Ltd., provider of genetic testing services in Russian Federation, participated in this study. Data were collected during 2019-2020 by an online survey on the Internet portal of Genotek Ltd. The study included respondents over 18 years old, who voluntarily gave their personal data. Respondents who did not complete the questionnaire were excluded from the study. Participants’ personal information was completely hidden during the study. All research was approved by the ethics committee of Genotek Ltd. The participants have provided written informed consent.

The questionnaire was in Russian and included three blocks. The first block contained questions about sex and age. The second block consisted of questions from the depression subscale of HADS (HADS-D). There were 7 given questions, each of which was offered 4 options of answer, assessed from 0 to 3 points depending on the severity of the symptom. The third block consisted of questions about whether the respondent had diagnoses of cardiovascular diseases (CVD) or certain symptoms associated with CVD. The respondents were asked to answer “yes” or “no” to the questions about the presence of the following conditions: “high blood pressure (hypertension)”, “cardiac rhythm disturbances”, “heart pain”, “shortness of breath”, “stroke”, “heart attack”, “lower extremity vessel thrombosis”, “pulmonary embolism”, “varicose veins”. The presence of at least one affirmative answer to the questions from this block was the reason for classifying the respondent as a person with possible CVD.

To identify the differences between the respondents depending on age, groups by age were formed based on the analysis of the age distribution in the studied cohort using quartiles (Q1 = 30, Q2 (Me) = 35, Q3 = 42). Accordingly, four age groups of patients were identified: up to 30 y.o., 31 to 35 y.o., 36 to 42 y.o. and 43 y.o. and older.

We analyzed the differences between groups for two characteristics associated with the HADS-D score for each participant: the mean score (quantitative assessment) and the category of the participant determined by the individual score: “no depression” (0-7 points), “subclinical depression” (8-10 points) and “clinical depression” (11 or more points) [10].

To analyze statistical data the R programming language was used [14]. For the analysis of quantitative variables, the nonparametric Wilcoxon-Mann-Whitney and the Kruskal-Wallis tests were used. The Fisher’s exact test was used for the analysis of nominal variables. If there were multiple comparisons, Bonferroni and Holm corrections were applied. Multiple linear regression was used to assess the influence of factors (sex, CVD and age) on the HADS-D score. The effects of factors on the risk of developing depressive symptoms (as determined by HADS-D), was assessed using binary logistic regression. The absence of depression by HADS-D (0-7 points) and the presence of depressive symptoms at the subclinical and clinical levels (8 or more points) were chosen as binary outcomes. It is worth noting that a meta-analysis of studies using HADS-D proved that 8 points are the optimal threshold value for detecting the presence of depressive symptoms in people without psychiatric diagnoses [11].

## RESULTS

### Sample characteristics

The final sample size was 2610 people, of which 51.7% were women (1349). The average age of the respondents was (M (SD)) 36.79 (9.67) years. The presence of CVDs was reported by 30.4% (794) of the respondents.

The overall results for HADS-D are shown in Table 1. Respondents with clinical depression were significantly younger compared to participants without depressive symptoms (M (SD): 34.5 (10.5) vs. 37.0 (9.7), p = 0.038). There were no age differences between participants with clinical and subclinical depression (36.0 (9.3)) (p = 0.342).

**Table 1.** Sample characteristics

|  |  |
| --- | --- |
| Age (M(SD)), years | 36,79 (9,67) |
| % of women (n) | 51,7% (1349). |
| % of respondents with CVD (n) | 30,4% (794) |
| *HADS-D results* |  |
| Score (M(SD)), points | 4,4 (2,8) |
| 0-7 points (no depression), % (n) | 85,7% (2236) |
| 8-10 points (subclinical depression), % (n) | 11,5% (299) |
| 11 and more points (clinical depression), % (n) | 2,9% (75) |
| Total (n) | 2610 |

### Gender differences

In women the mean HADS-D score was significantly higher (p=0.003; Table 2). Among women, the proportion of respondents with clinical depression (in comparison with the proportion of no-depression group) was higher compared to men (p=0.046), however, after applying the correction for multiple comparisons, this difference ceased to reach the level of statistical significance (p=0.139). Also, men were significantly older than women (p <0.0001). There were no significant differences in the incidence of CVD (p=0.202) between men and women. Thus, women, despite a higher mean score, did not differ from men in the incidence of clinical and subclinical depression as assessed by HADS-D.

**Table 2.** Sex differences

| Sex | Men | Women |
| --- | --- | --- |
| Age (M(SD)), years | **37,6 (9,7)\*** | 36,1 (9,5) |
| % of respondents with CVD (n) | 31,6% (399) | 29,3% (395) |
| *HADS-D results* |  |  |
| Score (M(SD)), points | 4,2 (2,7) | **4,6 (2,9)\*** |
| 0-7 points (no depression), % (n) | 87,15% (1099) | 84,29% (1137) |
| 8-10 points (subclinical depression), % (n) | 10,63% (134) | 12,23% (165) |
| 11 and more points (clinical depression), % (n) | 2,22% (28) | 3,48% (47) |
| Total (n) | 1261 | 1349 |

\* - $p < 0,01$ between sex groups

### Age differences

It was found that in the group of young respondents (up to 30 y.o.), the average HADS-D score was significantly higher compared to older respondents (43 and older) (p = 0.004; Table 3, Fig. 1), as well as compared with participants aged 36 to 42, although this difference is only a trend (p = 0.082).

**Table 3.**
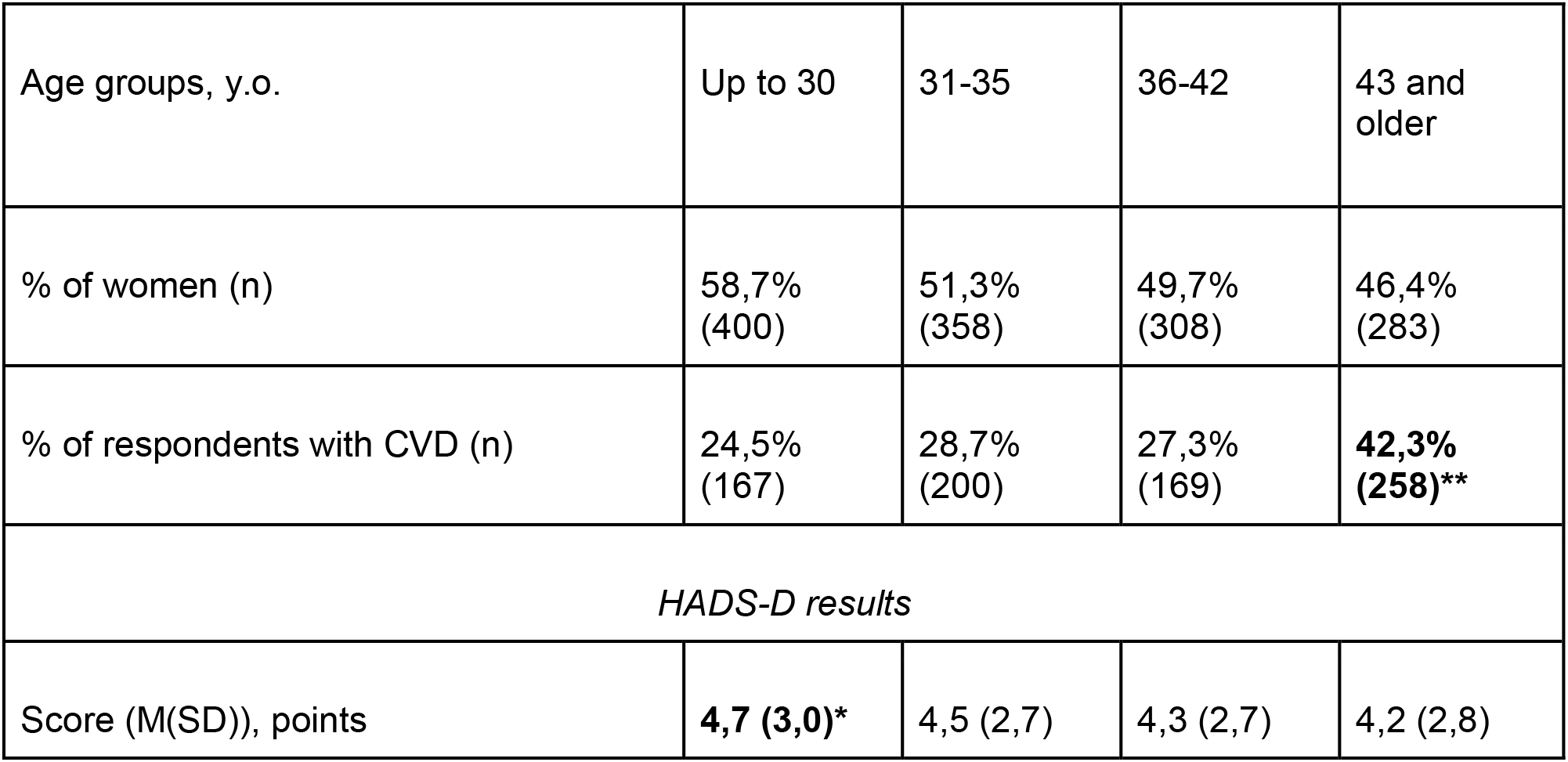
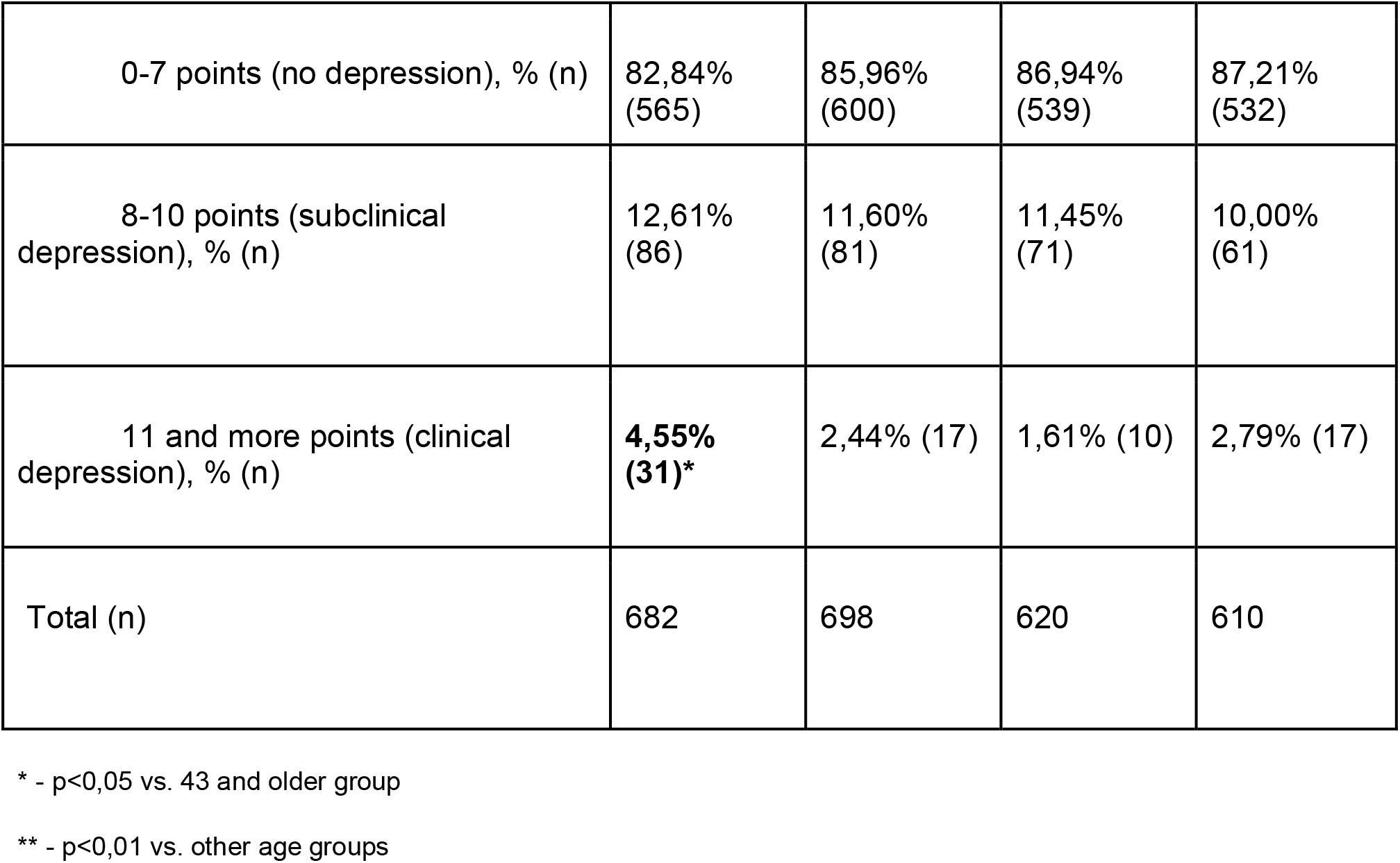
Age differences

| Age groups, y.o. | Up to 30 | 31-35 | 36-42 | 43 and older |
| --- | --- | --- | --- | --- |
| % of women (n) | 58,7%<br>(400) | 51,3%<br>(358) | 49,7%<br>(308) | 46,4%<br>(283) |
| % of respondents with CVD (n) | 24,5%<br>(167) | 28,7%<br>(200) | 27,3%<br>(169) | **42,3%**<br>**(258)\*\*** |
| *HADS-D results* |  |  |  |  |
| Score (M(SD)), points | **4,7 (3,0)\*** | 4,5 (2,7) | 4,3 (2,7) | 4,2 (2,8) |
| 0-7 points (no depression), % (n) | 82,84%<br>(565) | 85,96%<br>(600) | 86,94%<br>(539) | 87,21%<br>(532) |
| 8-10 points (subclinical depression), % (n) | 12,61%<br>(86) | 11,60%<br>(81) | 11,45%<br>(71) | 10,00%<br>(61) |
| 11 and more points (clinical depression), % (n) | **4,55%**<br>**(31)\*** | 2,44% (17) | 1,61% (10) | 2,79% (17) |
| Total (n) | 682 | 698 | 620 | 610 |

\* - $p < 0,05$ vs. 43 and older group
\*\* - $p < 0,01$ vs. other age groups

**Fig. 1.**
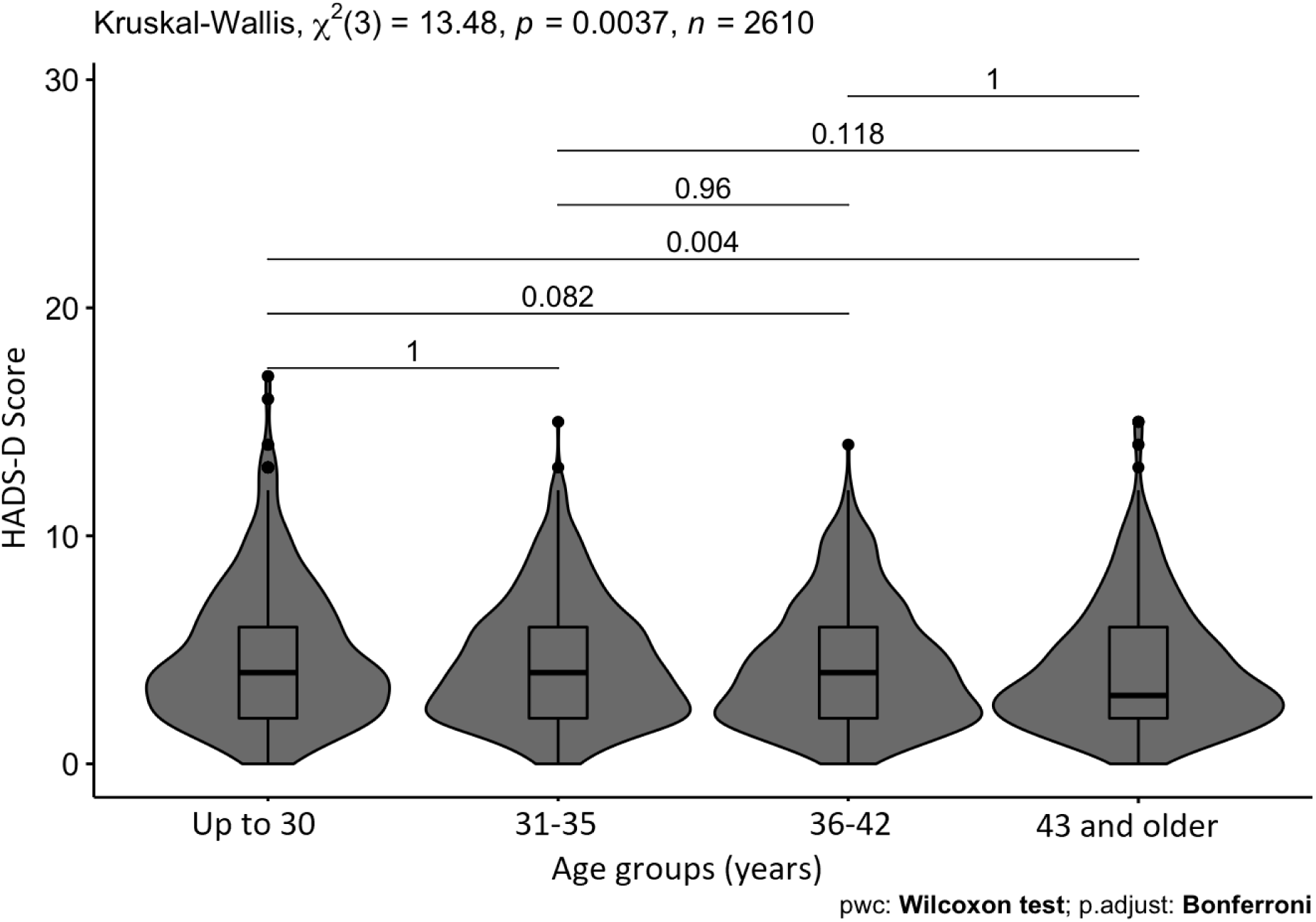
Box-plots and violin-plots of differences between age groups. The gray areas show the density of distribution of HADS-D scores in age groups.

In the group of young respondents up to 30 y.o., the proportion of participants with clinically significant depression (when compared with the proportion of participants without depressive symptoms) was significantly higher than in the group of respondents aged 36 to 42 (p = 0.00134). In addition, in the group of older respondents (43 and older), the proportion of respondents with CVD was significantly higher compared to other age groups (p <0.01)

### Differences between respondents with and without CVD

The mean HADS-D score among respondents with CVD was higher (p<0.0001; Table 4). However, in the group of respondents with CVD, the proportions of participants with subclinical and clinical depression did not differ significantly from those in the group without CVD. Groups with and without CVD did not differ in the proportion of women (p=0.202), however, participants with CVD were significantly older (p <0.0001).

**Table 4.** Differences between respondents with and without CVD

| CVD | Without CVD | With CVD |
| --- | --- | --- |
| Age (M(SD)), years | 35,8 (9,0) | **39,0 (10,8)\*** |
| % of women (n) | 52,5% (954) | 49,7% (395) |
| *HADS-D results* |  |  |
| Score (M(SD)), points | 4,3 (2,7) | **4,8 (2,8)\*\*** |
| 0-7 points (no depression), % (n) | 86,4% (1570) | 83,9% (666) |
| 8-10 points (subclinical depression), % (n) | 11,1% (201) | 12,3% (98) |
| 11 and more points (clinical depression), % (n) | 2,5% (45) | 3,8% (30) |
| Total (n) | 1816 | 794 |

\* - $p<0,05$ between CVD groups, \* - $p<0,01$ between CVD groups

### Regression analysis

To assess the possible role of sex, age and the presence of CVD as a risk factors for depression assessed by HADS-D, regression analysis was performed for the HADS-D score (linear regression) and for the risk of falling into the category of respondents with depressive symptoms, both at clinical and subclinical levels (binary logistic regression).

Linear regression showed that the presence of CVD leads to a significant increase in the HADS-D score (p <0.001, β = 0.57, 95% CI [0.33 - 0.80]). On the contrary, male sex decreases the HADS-D score (p=0.002, β =−0.33, 95% CI [−0.54 – −0.12]), higher age also decreases the HADS-D score (p<0.001, β =−0.02, 95% CI [−0.03 – −0.01]).

Logistic regression (Fig. 2) showed that the presence of CVD is a risk factor for the presence of symptoms of depression (p=0.033, OR=1.29). On the contrary, older age reduces the risk (p=0.015, OR=0.87) as a male sex too (trend, p=0.052, OR=0.80).

**Fig. 2.**
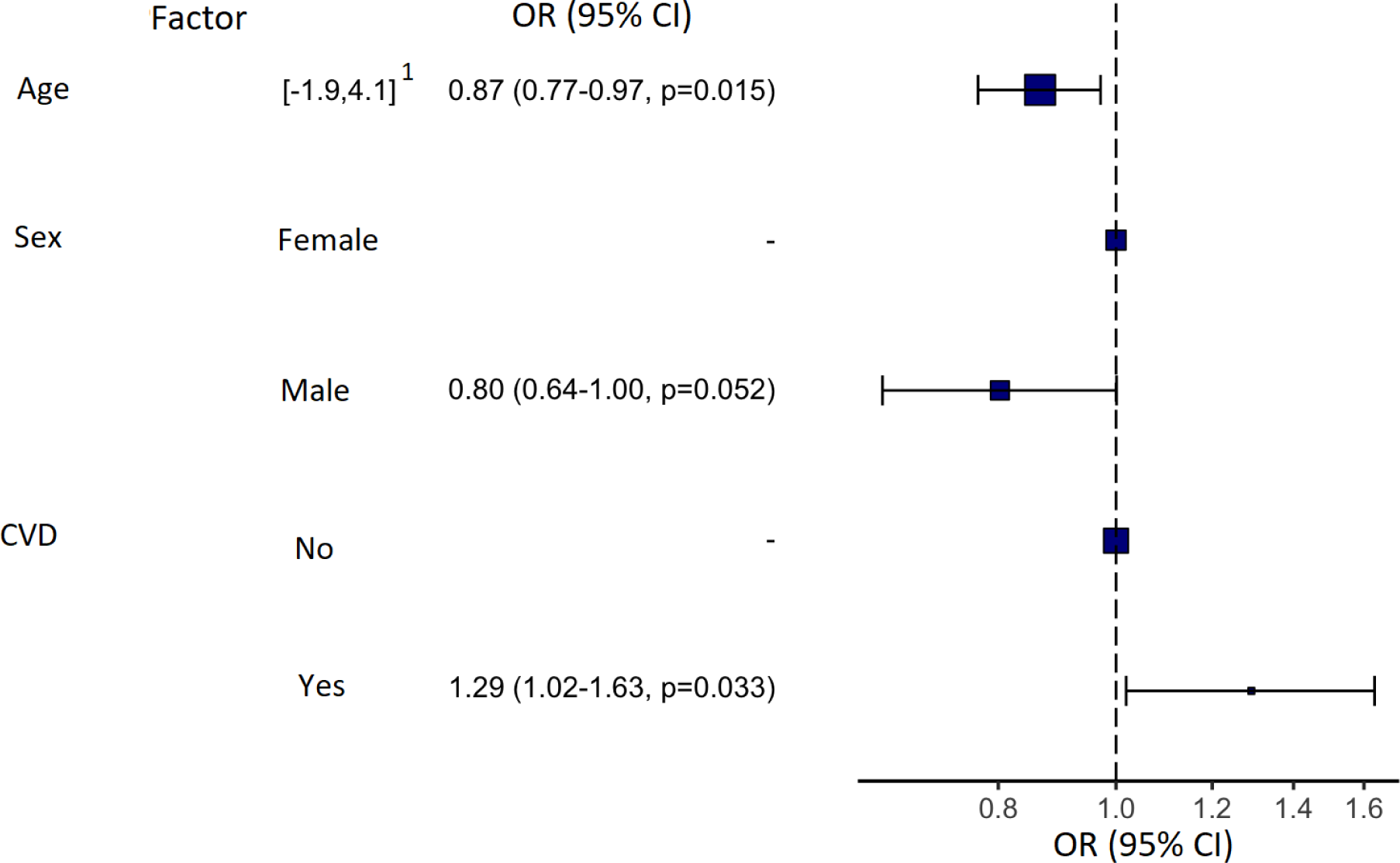
Results of binary logistic regression for assessing the risk of having symptoms of clinical or subclinical depression according to HADS-D. 1 - spread of values after normalization

## DISCUSSION

In our study the online version of HADS-D revealed the same population patterns found with other screening methods for depression. Thus, using this phenotyping method, we demonstrated that a greater severity of depressive symptoms is associated with the female sex, younger age and the presence of CVD.

According to the results we obtained using the online HADS-D questionnaire as a screening method for depression, the proportion of respondents with depressive symptoms was 14.4% (of which 11.5% were at subclinical level, and 2.9% – at clinical). Our data is somewhat lower than the results obtained by Shalnova et al. in the study of the prevalence of anxiety and depression in the general Russian population using HADS, conducted in 2012-2013 during the epidemiological study of CVD. It should be noted that HADS was used in a “paper” format and was a part of extensive medical examination with the use of a variety of laboratory and instrumental methods. In this study, the proportion of respondents with subclinical depression (HADS-D ≥ 8) was 25.6%; and with clinical depression (HADS-D≥11) – 8.8% [15]. However, in other studies using HADS, conducted on European populations, the proportion of respondents with 8 or more points was lower – from 11% to 23.5%; however, the proportion of patients with 11 or more points was comparable, ranging from 5% to 9.6% [16]-[19]. The mean HADS-D score in our study was 4.4 points, which is also comparable to the data on the mean HADS-D score in the European population, which ranged from 3.0 to 4.7 points [16], [17], [20] - [22].

In our study the mean HADS-D score was higher in women. This result is confirmed by numerous studies (also using self-questionnaires) indicating a greater severity of depression in women compared to men. In a study by Shalnova et al. there have been demonstrated similar differences in HADS-D scores between genders [15]. Gender differences in the severity of depression (assessed using Beck Depression Inventory (BDI) and General Health Questionnaire (GHQ) scales) were also demonstrated in the European population [23]. However, according to the results of our study, the proportion of respondents with depressive symptoms did not differ significantly between sex groups. Although data from epidemiological studies based on the formal diagnostic criteria of the DSM or ICD show a higher incidence of depression in women [24], some HADS studies indicate that there is no such difference. For example, a study by Hinz et al., conducted in a German population, demonstrated no significant differences between men and women in terms of both the mean score and the proportion of respondents with depressive disorders [17]. One of the largest HADS studies in the general population, conducted in Norway, also found no significant difference between sex groups in the proportion of respondents with depressive disorders [21]. In addition, some studies have found even higher rates of depression when assessed by HADS-D in men [18], [25]. Several explanations have been proposed for the observed “reverse” sex pattern of depression as measured by HADS-D. Some researchers argue that depression in women is more likely to manifest with somatic symptoms, therefore, the absence of questions about them in HADS-D leads to a decrease in the detection of depressive disorders in women [26]. Other authors demonstrate that this difference may be due to the fact that HADS-D questions are able to identify only one of the possible clinical subtypes of the disease - “anhedonic” depression [21]. At the same time, studies of anhedonia have shown the absence of sex differences, which, in turn, may cause changes in the sex structure of depression when assessed by HADS-D [25]. In addition, much of the epidemiological data on the higher incidence of depression in women is based primarily on treatment demand. Men are known to seek help less often and, thus, more cases of depression in men remain undiagnosed. It has been shown that treatment demand in men is associated with a greater severity of health problems than in women [27]. In our study design participants did not seek medical care for depression and that could also reduce the observed difference between men and women in depression. Moreover, this fact may be an advantage of our study, making it possible to identify a more objective picture of the distribution of depressive symptoms in the population outside of their connection with the use of medical care.

We have demonstrated that participant’s age also was associated with the severity and incidence of depressive symptoms as assessed by HADS-D. Depression has been considered more common in middle-aged and seniors and that was demonstrated in population studies conducted in the late 20th and early 21st centuries [24]. An increase in the incidence of depressive disorders in older age groups has also been demonstrated in various studies using HADS-D [17], [21]. However, our results indicate a greater severity of depressive symptoms in the young age group (up to 30 y.o.), which correlates with the results of newer studies. For example, in a 2012 study by Patten et al., conducted on Canadian adults, there was a 2% decrease in the annual prevalence of depression with each year of life [28]. Similar results were demonstrated in a study by Almas et al. conducted on a Swedish population. Authors found that depressed patients (assessed by Major Depression Inventory) were younger than healthy participants. [29]. These results can be explained by a significant increase in the incidence of depression in the young age group over the past 15 years. According to the results of the 2017 National Survey on Drug Use and Health study conducted in the United States, the annual prevalence of depression in the 18-25 age group was significantly higher compared to older groups [30].

Our results show that CVD, self-reported by respondents, was also a factor associated with the severity of depressive symptoms when assessed by online HADS-D. Although the proportion of patients with depressive symptoms did not differ significantly among respondents with CVD compared with healthy respondents, logistic regression analysis showed that the presence of CVD increases the risk of developing depressive symptoms (OR = 1.29). These results are consistent with the known data on a higher level of depression in patients with CVD in various populations, including Russian one [15], [24].

### Strengths

To our knowledge, this is the first study with the evaluation of the online version of HADS-D for phenotyping depression in the general population. A large sample of respondents was studied, and positive results were obtained, which makes it possible to use this tool as one of the methods of phenotyping depression in the general population. New data were obtained on the prevalence of depressive symptoms in the Russian population, as well as on the association of sex, age, and the presence of cardiovascular diseases on the severity of depressive symptoms.

### Limitations

This study had several limitations. We did not use diagnostic tools to assess prevalence of depression as a clinical diagnosis in our sample and were unable to compare these rates with the results of screening with HADS-D. In addition, HADS-D allows identifying only depressive symptoms that are relevant at the time of testing and does not allow assessing the possible presence of depression in the past. Also, we did not conduct medical examinations of respondents and did not have access to their medical records, and therefore we determined the presence of CVD solely on the basis of self-report.

## CONCLUSIONS

Online HADS-D demonstrated results similar to those obtained with the “paper” version, which assumes personal contact between respondents and researchers. In addition, the population patterns of depression prevalence identified with the online version of HADS-D are consistent with the results of large-scale epidemiological studies of depression. Thus, the online HADS-D, as a simpler and less expensive method with satisfactory applicability and may be efficient enough for depression phenotype screening in the general population. However, further studies on larger samples are needed to more accurately determine the capabilities and limitations of this tool in depression phenotyping for genetic studies.

## Data Availability

STROBE (EQUATOR Network) check-list for this study: https://dfiles.eu/files/jqd7cqtro?redirect

## Notes

**Funding:** This research was supported by Russian Science Foundation Grant №20-15-00132

### Competing Interest Statement

The authors have declared no competing interest.

### Funding Statement

This research was supported by Russian Science Foundation Grant №20-15-00132

### Author Declarations

All research was approved by the ethics committee of Genotek Ltd.

## REFERENCES

1. WHO, “Depression - Fact Sheet,” 2018. https://www.who.int/news-room/fact-sheets/detail/depression.

2. Levinson DF, Mostafavi S, Milaneschi Y, Rivera M, Ripke S, Wray NR, et al. Genetic Studies of Major Depressive Disorder: Why Are There No Genome-wide Association Study Findings and What Can We Do About It? Biol Psychiatry 2014; 76: 510–2. https://doi.org/10.1016/j.biopsych.2014.07.029.

3. Smoller JW, Andreassen OA, Edenberg HJ, Faraone S V., Glatt SJ, Kendler KS. Psychiatric genetics and the structure of psychopathology. Mol Psychiatry 2019; 24 :409–20. https://doi.org/10.1038/s41380-017-0010-4.

4. Schwabe I, Milaneschi Y, Gerring Z, Sullivan PF, Schulte E, Suppli NP, et al. Unraveling the genetic architecture of major depressive disorder: merits and pitfalls of the approaches used in genome-wide association studies. Psychol Med 2019; 49: 2646–56. https://doi.org/10.1017/S0033291719002502.

5. Ahmed AT, Frye MA, Rush AJ, Biernacka JM, Craighead WE, McDonald WM, et al. Mapping depression rating scale phenotypes onto research domain criteria (RDoC) to inform biological research in mood disorders. J Affect Disord 2018; 238:1–7. https://doi.org/10.1016/j.jad.2018.05.005.

6. Cai N, Revez JA, Adams MJ, Andlauer TFM, Breen G, Byrne EM, et al. Minimal phenotyping yields genome-wide association signals of low specificity for major depression. Nat Genet 2020; 52:437–47. https://doi.org/10.1038/s41588-020-0594-5.

7. Eid RS, Gobinath AR, Galea LAM. Sex differences in depression: Insights from clinical and preclinical studies. Prog Neurobiol 2019; 176:86–102. https://doi.org/10.1016/j.pneurobio.2019.01.006.

8. Weinberger AH, Gbedemah M, Martinez AM, Nash D, Galea S, Goodwin RD. Trends in depression prevalence in the USA from 2005 to 2015: Widening disparities in vulnerable groups. Psychol Med 2018; 48:1308–15. https://doi.org/10.1017/S0033291717002781.

9. Huffman JC, Celano CM, Beach SR, Motiwala SR, Januzzi JL. Depression and Cardiac Disease: Epidemiology, Mechanisms, and Diagnosis. Cardiovasc Psychiatry Neurol 2013; 2013:1–14. https://doi.org/10.1155/2013/695925.

10. Zigmond AS, Snaith RP. The Hospital Anxiety and Depression Scale. Acta Psychiatr Scand 1983; 67:361–70. https://doi.org/10.1111/j.1600-0447.1983.tb09716.x.

11. Brennan C, Worrall-Davies A, McMillan D, Gilbody S, House A. The Hospital Anxiety and Depression Scale: A diagnostic meta-analysis of case-finding ability. J Psychosom Res 2010; 69:371–8. https://doi.org/10.1016/j.jpsychores.2010.04.006.

12. Cronly J, Duff AJ, Riekert KA, Perry IJ, Fitzgerald AP, Horgan A, et al. Online versus paper-based screening for depression and anxiety in adults with cystic fibrosis in Ireland: a cross-sectional exploratory study. BMJ Open 2018; 8:e019305. https://doi.org/10.1136/bmjopen-2017-019305.

13. Andersson G, Kaldo-Sandström V, Ström L, Strömgren T. Internet administration of the Hospital Anxiety and Depression Scale in a sample of tinnitus patients. J Psychosom Res 2003; 55:259–62. https://doi.org/10.1016/S0022-3999(02)00575-5.

14. R Core team. R: A language and environment for statistical computing. R Foundation for Statistical Computing, Vienna, Austria 2020. https://www.r-project.org/.

15. Shalnova SA, Evstifeeva SE, Deev AD, Artamonova GV, Gatagonova TM. [The prevalence of anxiety and depression in different regions of the Russian Federation and its association with sociodemographic factors (according to the data of the ESSE-RF study)]. Article in Russian.Ter. Arkh., vol. 86, no. 12, pp. 53–60, 2014, doi: 10.17116/terarkh2014861253-60.

16. Lisspers J, Nygren A, Söderman E. Hospital anxiety and depression scale (HAD): Some psychometric data for a Swedish sample. Acta Psychiatr Scand 1997;96:281–6. https://doi.org/10.1111/j.1600-0447.1997.tb10164.x.

17. Hinz A, Brähler E. Normative values for the hospital anxiety and depression scale (hads) in the general german population. J Psychosom Res 2011;71:74–8. https://doi.org/10.1016/j.jpsychores.2011.01.005.

18. Nortvedt MW, Riise T, Sanne B. Are men more depressed than women in Norway? Validity of the Hospital Anxiety and Depression Scale. J Psychosom Res 2006;60:195–8. https://doi.org/10.1016/j.jpsychores.2005.07.002.

19. Breeman S, Cotton S, Fielding S, Jones GT. Normative data for the Hospital Anxiety and Depression Scale. Qual Life Res 2015;24:391–8. https://doi.org/10.1007/s11136-014-0763-z.

20. Crawford JR, Garthwaite PH, Lawrie CJ, Henry JD, MacDonald MA, Sutherland J, et al. A convenient method of obtaining percentile norms and accompanying interval estimates for self-report mood scales (DASS, DASS-21, HADS, PANAS, and sAD). Br J Clin Psychol 2009;48:163–80. https://doi.org/10.1348/014466508X377757.

21. Stordal E, Bjartveit Krüger M, Dahl NH, Krüger O, Mykletun A, Dahl AA. Depression in relation to age and gender in the general population: The Nord-Trøndelag health study (HUNT). Acta Psychiatr Scand 2001;104:210–6. https://doi.org/10.1034/j.1600-0447.2001.00130.x.

22. Spinhoven P, Ormel J, Sloekers PPA, Kempen GIJM, Speckens AEM, Van Hemert AM. A validation study of the hospital anxiety and depression scale (HADS) in different groups of Dutch subjects. Psychol Med 1997;27:363–70. https://doi.org/10.1017/S0033291796004382.

23. Aalto A-M, Elovainio M, Kivimäki M, Uutela A, Pirkola S. The Beck Depression Inventory and General Health Questionnaire as measures of depression in the general population: A validation study using the Composite International Diagnostic Interview as the gold standard. Psychiatry Res 2012;197:163–71. https://doi.org/10.1016/j.psychres.2011.09.008.

24. Gutiérrez-Rojas L, Porras-Segovia A, Dunne H, Andrade-González N, Cervilla JA. Prevalence and correlates of major depressive disorder: a systematic review. Brazilian J Psychiatry 2020;00. https://doi.org/10.1590/1516-4446-2020-0650.

25. Langvik E, Hjemdal O, Nordahl HM. Personality traits, gender differences and symptoms of anhedonia: What does the Hospital Anxiety and Depression Scale (HADS) measure in nonclinical settings?Scand J Psychol 2016;57:144–51. https://doi.org/10.1111/sjop.12272.

26. Piccinelli M, Wilkinson G. Gender differences in depression. Br J Psychiatry 2000;177:486–92. https://doi.org/10.1192/bjp.177.6.486.

27. Call JB, Shafer K. Gendered Manifestations of Depression and Help Seeking Among Men. Am J Mens Health 2018;12:41–51. https://doi.org/10.1177/1557988315623993.

28. Patten SB, Williams JVA, Lavorato DH, Wang JL, McDonald K, Bulloch AGM. Descriptive epidemiology of major depressive disorder in Canada in 2012. Can J Psychiatry 2015;60:23–30. https://doi.org/10.1177/070674371506000106.

29. Almas A, Forsell Y, Iqbal R, Janszky I, Moller J. Severity of depression, anxious distress and the risk of cardiovascular disease in a Swedish population-based cohort. PLoS One 2015;10:1–12. https://doi.org/10.1371/journal.pone.0140742.

30. US NIMH. Major Depression. Prevalence of Major Depressive Episode Among Adults. 2019. https://www.nimh.nih.gov/health/statistics/major-depression.shtml.

